# Comparative Evaluation Of Machine Learning Classifiers For Brain Tumor Detection

**DOI:** 10.1101/2024.07.28.24311114

**Authors:** Umair Ali

## Abstract

This study evaluates the effectiveness of six machine learning classifiers—Support Vector Classifier (SVC), Logistic Regression, K-Nearest Neighbors (KNN), Naive Bayes, Decision Tree, and Random Forest—in detecting brain tumors using numerical data rather than traditional imaging techniques like MRI. The results emphasize the importance of data preprocessing, particularly feature scaling, in enhancing model performance. Among the classifiers, Random Forest emerged as the top performer, achieving an accuracy of 98.27% on both original and scaled data, demonstrating its robustness and reliability. The study highlights the potential of Random Forest as a valuable tool for automated brain tumor detection in clinical settings, offering a cost-effective and accessible alternative for resource-constrained environments. The paper suggests that future research should explore advanced deep learning models, such as 3D Convolutional Neural Networks (CNNs) and Generative Adversarial Networks (GANs), to further improve diagnostic accuracy and support early intervention and personalized treatment strategies for brain tumor patients.

## I. Introduction

Brain tumors represent a significant global health concern, with the Nature Brain Tumor Society (NBTS) estimating that approximately 700,000 individuals in the United States are affected by these malignancies each year [1]. Brain tumors can be classified into various categories such as gliomas, medulloblastomas, and acoustic neuromas, each presenting distinct characteristics and treatment challenges [2]. Effective early detection and accurate diagnosis of brain tumors are crucial for optimizing treatment strategies and improving patient outcomes. Untreated brain tumors can lead to severe neurological deficits, cognitive impairments, and, in many cases, death [3].

Traditionally, brain tumors have been diagnosed using Magnetic Resonance Imaging (MRI), which offers high-resolution images of brain structures [4]. However, analyzing MRI data manually is both time-consuming and prone to variability, and often lacks the precision required for accurate tumor detection and segmentation [5]. This challenge has spurred interest in leveraging machine learning techniques to automate and enhance the diagnostic process.

Recent advancements in machine learning and artificial intelligence have shown promising potential in the realm of medical diagnostics [6]. In particular, machine learning models that rely on numerical values extracted from patient data, such as clinical features, genetic information, and laboratory results, have been increasingly explored as a means of improving brain tumor detection [7]. These models can offer significant advantages over traditional image-based methods by facilitating faster and more consistent diagnostic processes [3].

This study aims to evaluate the effectiveness of several well-known machine learning classifiers for the task of brain tumor detection using numerical data. Specifically, we examine the performance of Support Vector Classifier (SVC), Logistic Regression Classifier, K-Nearest Neighbors (KNN) Classifier, Naive Bayes Classifier, Decision Tree Classifier, and Random Forest Classifier. Each of these algorithms brings unique strengths to the table. For instance, SVM is known for its effectiveness in high-dimensional spaces and its ability to handle non-linearly separable data [8]. Logistic Regression is appreciated for its simplicity, interpretability, and capability to manage both continuous and categorical features [9]. KNN is valued for its robustness to noise and ability to capture complex feature interactions [10]. Naive Bayes offers benefits in handling categorical data and learning from smaller datasets [11]. Decision Trees are favored for their interpretability and ability to model both categorical and numerical features [12]. Random Forest, an ensemble method, is known for reducing overfitting and handling high-dimensional data effectively [13].

The motivation behind using these classifiers lies in their distinct advantages for processing numerical data and their varying approaches to handling complex patterns in the data. This study leverages a dataset consisting of clinical and diagnostic numerical values related to brain tumors, providing a platform for evaluating the performance of these classifiers [14]. The aim is to determine which classifier provides the highest accuracy and reliability for brain tumor detection, contributing to the development of efficient diagnostic tools [15].

Machine learning has demonstrated significant promise in the medical field, with various studies highlighting its effectiveness in improving diagnostic accuracy [16]. For instance, recent research has shown that machine learning models can significantly enhance the accuracy of cancer detection and prognosis prediction [17]. By applying these techniques to brain tumor detection using numerical data, this study seeks to build upon these advancements and offer a novel approach to diagnosing brain tumors [18].

In this study, this study focuses on evaluating the efficacy of various machine learning classifiers in detecting brain tumors from numerical values rather than MRI images. The goal is to identify the most effective algorithm for this task, thereby contributing to the broader effort of improving brain tumor diagnosis and ultimately enhancing patient outcomes.

The paper is further structured as follows: Section II discusses the literature review on the brain tumor detection. Section III highlights motivation. Section IV describes machine learning algorithms. Section V presents methodology. Section VI result. Section VII discussion. Lastly, Section VIII concludes the paper and future work.

## II. Literature Review

This paper [19] highlights the “curse of dimensionality” often encountered in brain tumor datasets with many features. They propose a two-pronged approach: first, using Particle Swarm Optimization to select the most informative features, mimicking the efficient foraging behavior of birds or fish. Second, they employ ensemble learning with Majority Voting, combining the predictions of multiple classifiers to improve accuracy and robustness. This approach could be particularly relevant to your work if you’re dealing with a large number of features.

While the paper [20] utilizes an SVM, its core contribution lies in a novel feature extraction method designed to capture the most discriminative information from brain tumor data. This emphasis on feature engineering is highly transferable. You could apply their proposed feature extraction techniques and then experiment with alternative classifiers like Random Forest, which is known for its ability to handle high-dimensional data, or Gradient Boosting, which excels at reducing bias and achieving high accuracy.

This paper [21] delves into the realm of unsupervised learning for brain tumor detection, specifically employing the K-Means clustering algorithm. K-Means groups similar data points together based on their features, aiming to uncover hidden patterns and structures within the data without relying on labeled examples. This approach could be beneficial for your research by potentially revealing distinct clusters or subgroups within your dataset that correspond to different tumor characteristics or stages.

While the [22] title mentions CNNs, which are typically used for image data, this paper emphasizes the critical role of feature extraction for accurate brain tumor classification, regardless of the data type. They highlight how carefully engineered features can significantly improve the performance of machine learning models. You can draw inspiration from their feature engineering techniques and apply them to your tabular data to potentially enhance the accuracy of your chosen classifiers.

[23] Brain tumor detection and segmentation have been extensively explored using machine learning and deep learning techniques. Various studies have proposed CNN-based methods, automated feature extraction and classification approaches, and comparisons of deep learning models. Additionally, hybrid approaches combining different techniques have been investigated. These studies have achieved high accuracy rates, ranging from 91.43% to 98.69%, demonstrating the potential of machine learning and deep learning in brain tumor detection and segmentation.

[24] Brain tumor segmentation has been extensively explored using machine learning techniques. Previous reviews have focused on traditional computer vision methods and deep learning approaches. Recent studies have investigated the use of convolutional neural networks (CNNs) for brain tumor segmentation. Other approaches include using transfer learning, ensemble learning, and hybrid models combining CNNs with traditional machine learning techniques. These studies demonstrate the potential of machine learning for brain tumor segmentation, achieving high accuracy and efficiency.

In study [25] MRI-based brain tumor detection using convolutional deep learning methods and machine learning techniques was explored. A 2D CNN and auto-encoder network were proposed, achieving training accuracies of 96.47% and 95.63%, respectively. Six machine learning techniques were compared, with KNN achieving the highest accuracy (86%) and MLP the lowest (28%). The study demonstrates the effectiveness of deep learning methods in brain tumor detection, with the proposed 2D CNN showing optimal accuracy and performance. This work contributes to the development of automated brain tumor detection systems, improving diagnosis and treatment.

## III. Motivation

Brain tumors are a leading cause of cancer-related deaths worldwide, with high mortality rates and a profound impact on the quality of life for patients and their families. Early and accurate diagnosis is crucial for effective treatment, improved patient outcomes, and enhanced survival rates. However, brain tumor diagnosis remains a challenging task, particularly in resource-constrained settings where access to advanced medical facilities, specialized personnel, and cutting-edge technologies is limited. In such settings, the lack of resources hinders the widespread adoption of advanced medical imaging techniques like MRI and CT scans, which are essential for accurate brain tumor diagnosis.

This research is motivated by the need for a cost-effective, objective, and accessible tool for brain tumor diagnosis that can operate within the constraints of resource-constrained settings. We aim to develop a predictive model that can aid in brain tumor diagnosis using readily available patient attributes and clinical features, eliminating the reliance on advanced medical imaging techniques or deep learning features. By leveraging machine learning algorithms and data analytics, our model seeks to provide a valuable tool for healthcare professionals, enabling them to make informed decisions and improve patient outcomes. Ultimately, our research strives to contribute to the development of efficient and accurate automated systems for early brain tumor diagnosis, leading to better patient care and treatment efficacy.

## IV. Machine Learning Classifiers

### A. Support Vector Classifier

Support Vector Machines are powerful supervised learning models used for classification and regression tasks. In the context of classification, an SVM aims to find an optimal hyperplane that best separates data points belonging to different classes.

The hyperplane is chosen to maximize the margin, which is the distance between the hyperplane and the closest data points from each class, known as support vectors. This focus on maximizing the margin contributes to the SVC’s ability to generalize well to unseen data [26].

SVCs can be applied to both linearly separable and non-linearly separable data. For non-linearly separable data, SVCs utilize kernel functions to map the data into a higher-dimensional space where it becomes linearly separable [27].

### B. Logistic Regression Classifier

Logistic regression is a statistical model used to predict the probability of a binary outcome (yes/no, 1/0) based on one or more independent variables. Unlike linear regression, which predicts continuous outcomes, logistic regression employs a sigmoid function to map predictions to a probability range between 0 and 1 [28].

This algorithm works by estimating the log odds of the outcome occurring based on the values of the independent variables. These log odds are then transformed into probabilities using the sigmoid function.

While primarily used for binary classification, logistic regression can be extended to handle multinomial outcomes (multiple categories) through variations like multinomial logistic regression. Logistic regression models are widely used in various fields, such as biology and social sciences, where the objective is to predict a categorical outcome [29].

### C. K-Nearest Neighbor (KNN) Classifier

The K-Nearest Neighbors (KNN) classifier is a straightforward yet powerful supervised learning algorithm used for both classification and regression tasks [30]. At its core, KNN operates on the principle that similar data points tend to cluster together.

When classifying a new data point, the algorithm identifies the *k* nearest neighbors to the point in the feature space, based on a chosen distance metric (e.g., Euclidean distance). The class label of the new data point is then determined by a majority vote among its *k* neighbors. For instance, if *k* is set to 5, and 3 out of the 5 nearest neighbors belong to class A, the new data point would be classified as belonging to class A.

One of the key advantages of KNN is its simplicity and ease of implementation [31]. It’s a non-parametric method, meaning it makes no assumptions about the underlying data distribution, making it suitable for datasets with complex or unknown structures. However, the choice of *k* is crucial, as a small *k* can make the model susceptible to noise, while a large *k* might lead to over smoothing and misclassification.

### D. Navie Bayes Classifier

The Naive Bayes Classifier (NBC) is a widely used machine learning algorithm for classification tasks. It is based on Bayes’ theorem, which describes the probability of a hypothesis given some observed evidence. In the context of classification, the hypothesis is the class label, and the evidence is the feature values of the instance to be classified [32].

The NBC algorithm assumes independence between features, meaning that each feature contributes independently to the probability of the class label. This assumption simplifies the calculation of the posterior probability of the class given the features. The algorithm calculates the likelihood of each feature given the class, as well as the prior probability of each class. Then, it applies Bayes’ theorem to calculate the posterior probability of each class given the features [32].

There are three main types of NBC, each suited to different types of data. Multinomial Naive Bayes (MNB) is used for multi-class problems with discrete features. Bernoulli Naive Bayes (BNB) is used for binary classification with binary features. Gaussian Naive Bayes (GNB) is used for continuous features and assumes a Gaussian distribution [33].

### E. Decision Tree Classifier

Decision Tree Classifiers are a popular supervised learning method used in machine learning for both classification and regression tasks [34]. Their strength lies in their intuitive, tree-like structure that breaks down complex decisions into a series of simpler ones, mirroring human-like reasoning. This makes them easy to understand and interpret, even for non-experts.

The algorithm works by recursively partitioning the dataset into increasingly homogeneous subsets based on the values of input features [35]. Starting at the root node, which represents the entire dataset, the algorithm searches for the best feature to split the data, aiming to create subsets that are as pure as possible in terms of class distribution [36]. This process continues down the tree, with each internal node representing a decision point based on a specific feature. The branches stemming from these nodes represent decision rules, guiding the data towards leaf nodes, which hold the final predictions or class labels.

### F. Random Forest Classifier

The Random Forest Classifier is a powerful ensemble learning method used in machine learning for both classification and regression tasks [37]. It operates by constructing a multitude of decision trees during training and outputting the class that is the mode of the classes (classification) or mean/average prediction (regression) of the individual trees [38].

The “random” aspect of Random Forest stems from two key concepts: random sampling of the training data and random subspace selection. During the creation of each tree, a technique called bootstrap sampling is employed, where the algorithm randomly selects a subset of the training data with replacement [39]. This means that some data points may be selected multiple times, while others might be left out. This process introduces diversity among the trees, as each tree learns from a slightly different perspective of the data.

## V. Methodology

### A. Introduction

The goal of this study is to develop and evaluate machine learning models to detect brain tumors using a dataset containing numerical values rather than images. This study employs multiple Machine Learning Classifiers, including Support Vector Classifier (SVC), Logistic Regression Classifier, K-Nearest Neighbors (KNN) Classifier, Naive Bayes Classifier, Decision Tree Classifier, and Random Forest Classifier, to classify individuals as having brain tumors or being healthy. The following sections detail the comprehensive methodology implemented in this study, which is divided into data preprocessing, model building, evaluation, and validation steps.

### B. Data Collection

The dataset used in this study consists of MRI images of brain tumors sourced from [40] [41]. These images were preprocessed to extract numerical features relevant to brain tumor detection, resulting in a dataset with 3761 rows and multiple columns. The columns include ‘Class’ (indicating the presence or absence of a brain tumor), as well as various texture features such as ‘Mean’, ‘Variance’, ‘Standard Deviation’, ‘Entropy’, ‘Skewness’, ‘Kurtosis’, ‘Contrast’, ‘Energy’, ‘ASM’, ‘Homogeneity’, ‘Dissimilarity’, and ‘Correlation’.

Sample images from the dataset are shown in Figure 1, which displays four MRI images (fig 1(a), fig 1(b), fig 1(c), fig 4(d)) of patients without brain tumors, while Figure 2 displays four MRI images (fig 2(a), fig 2(b), fig 2(c), fig 2(d)) of patients with brain tumors.

**Fig. 1.**
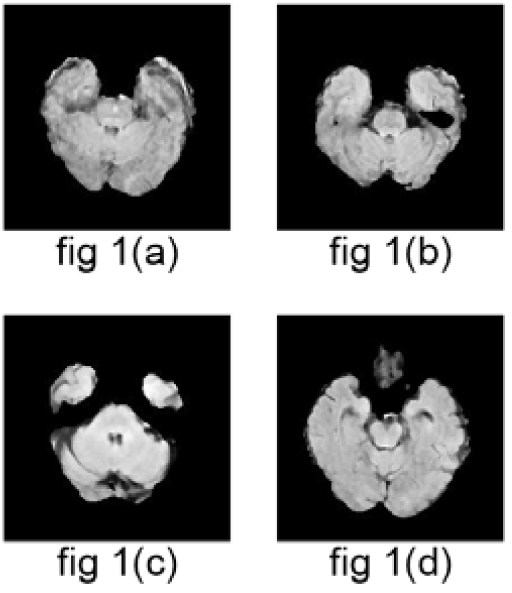
MRI images of patients without brain tumor.

**Fig. 2.**
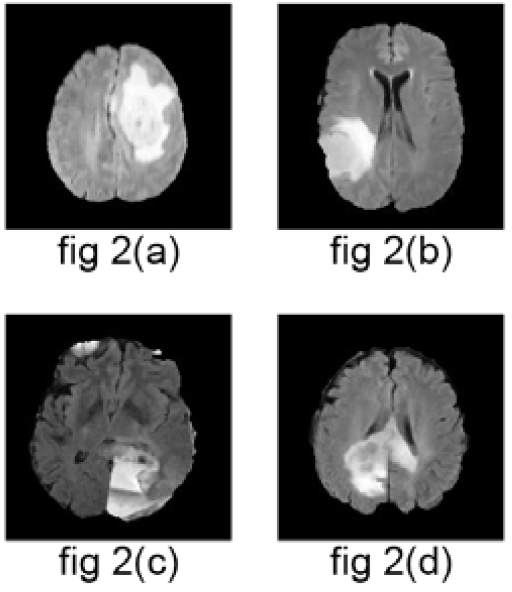
MRI images of patients with brain tumor.

### C. Preprocessing Steps

The extracted features were preprocessed to ensure data quality and consistency. The preprocessing steps included normalization, noise reduction, and feature scaling.

#### a) Import Libraries

First, essential libraries were imported for data analysis, visualization, and model. As shown in Table 1, the necessary libraries were imported using the following code:

**TABLE I.** Importing Libraries for data analysis and visualization.

| Code | Description |
| --- | --- |
| import numpy as np | Import NumPy library for numerical computations |
| import pandas as pd | Import Pandas library for data manipulation and analysis |
| import matplotlib.pyplot as plt | Import Matplotlib library for data visualization |
| import seaborn as sns | Import Seaborn library for data visualization and exploration |

#### b) Load Dataset

The dataset was loaded into a pandas DataFrame for analysis, as shown in Table 2.

**TABLE II.**
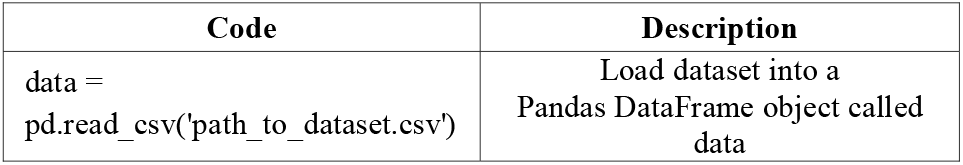
Loading dataset into pandas dataframe.

| Code | Description |
| --- | --- |
| data =<br>pd.read_csv('path_to_dataset.csv') | Load dataset into a Pandas DataFrame object called data |

**TABLE III.** Data Overview.

| Code | Description |
| --- | --- |
| data.info() | Summary of data structure |
| data.isnull().sum() | Count of missing values |
| data.describe() | Statistical summary of data |

#### c) Data Overview

Display information and summary statistics of the dataset, shown in Table 3

### D. Exploratory Data Analysis (EDA)

EDA was performed to understand the distribution and characteristics of the dataset. Techniques such as histogram plotting, box plots, and scatter plots were used to visualize data trends and outliers. These visualizations helped in identifying potential issues such as class imbalance and guided the data preprocessing steps.

#### a) Heatmap

Heatmaps are a powerful visualization tool in machine learning, used to represent complex data insights intuitively. By leveraging heatmaps, machine learning practitioners can gain a deeper understanding of their data, develop more effective models, and communicate insights more effectively. The code used to generate the heatmap is presented in Table 4:

**TABLE IV.** Heatmap.

| Code | Description |
| --- | --- |
| plt.figure(figsize=(16,9)) | Set figure size (16x9 inches) for plotting |
| sns.heatmap(data) | Visualize data as a heatmap |

#### b) Heatmap of Correlation Matrix

The dataset was loaded into a pandas DataFrame for analysis, as shown in Table 5, and 6.

**TABLE V.** Correlation matrix.

| Code | Description |
| --- | --- |
| data.corr() | Calculate correlation matrix of the dataset |

**TABLE VI.** Heatmap of Correlation matrix.

| Code | Description |
| --- | --- |
| plt.figure(figsize=(10,8)) | Set figure size (10x8 inches) for plotting |
| sns.heatmap(data.corr(), annot = True, cmap = 'coolwarm', linewidths=2) | Visualize correlation matrix as a heatmap with annotations, coolwarm color scheme, and defined line widths |

### E. Data Splitting

Data splitting is a crucial step in machine learning, where the available data is divided into two subsets: training data and testing data. The training data is used to train the model, while the testing data is used to evaluate its performance. The code used to split the data into training and testing is presented in Table 7.

**TABLE VII.** Data splitting into training and testing sets.

| Code | Description |
| --- | --- |
| from sklearn.model_selection<br>import train_test_split<br>X = data.drop(['Class'], axis=1) | Import train_test_split function for splitting data<br>Separate features (X) by dropping the target column ('Class') |
| y = data['Class'] | Separate target variable (y) as the 'Class' column |
| X_train, X_test, y_train, y_test =<br>train_test_split(X, y, test_size =<br>0.2, random_state= 5) | Split data into training (80%) and testing sets (20%) with a fixed random seed |

### F. Feature Scaling

Feature scaling, also known as data normalization. It involves transforming numeric features into a common scale, usually between 0 and 1, to prevent differences in scales from affecting model performance. The code used for feature scaling is presented in Table 8.

**TABLE VIII.** Feature Scaling.

| Code | Description |
| --- | --- |
| from sklearn.preprocessing<br>import StandardScaler | Import StandardScaler class for data scaling |
| sc = StandardScaler() | Initialize a StandardScaler object for scaling data |
| X_train_sc =<br>sc.fit_transform(X_train) | Scale training data (X_train) and fit scaler to it |
| X_test_sc = sc.transform(X_test) | Scale testing data (X_test) using the same scaler |

### G. Model Building and Training

After preprocessing the data, various machine learning models were developed and trained to predict the target variable. Both the original and scaled datasets were used to train the models, allowing for a comprehensive evaluation of their performance.

#### a) Support Vector Classifier

The SVC was trained with both the original and scaled data. Training the model with the original data is presented in Table 9, and training with scaled data is presented in Table 10.

**TABLE IX.** Support Vector Classifier Trained with Original Data.

| Code | Description |
| --- | --- |
| from sklearn.svm import SVC | Import Support Vector Classifier (SVC) from scikit-learn |
| svc_classifier = SVC() | Initialize an SVC object with default parameters |
| svc_classifier.fit(X_train, y_train) | Train the SVC model on the training data (X_train, y_train) |
| y_pred_svc =<br>svc_classifier.predict(X_test) | Use the trained model to predict labels for the testing data (X_test) |
| accuracy_score(y_test,<br>y_pred_svc) | Calculate the accuracy of the model by comparing predicted labels (y_pred_svc) with actual labels (y_test) |

**TABLE X.** Support Vector Classifier Trained with Scaled Data.

| Code | Description |
| --- | --- |
| svc_classifier_sc = SVC() | Initialize another SVC object for scaled data |
| svc_classifier_sc.fit(X_train_sc,<br>y_train) | Train the SVC model on scaled training data (X_train_sc, y_train) |
| y_pred_svc_sc =<br>svc_classifier_sc.predict(X_test_sc) | Use the trained model to predict labels for scaled testing data (X_test_sc) |
| accuracy_score(y_test,<br>y_pred_svc_sc) | Calculate the accuracy of the model on scaled data by comparing predicted labels (y_pred_svc_sc) with actual labels (y_test) |

#### b) Logistic Regression Classifier

The LRC was trained with both the original and scaled data. Training the model with the original data is presented in Table 11, and training with scaled data is presented in Table 12.

**TABLE XI.**
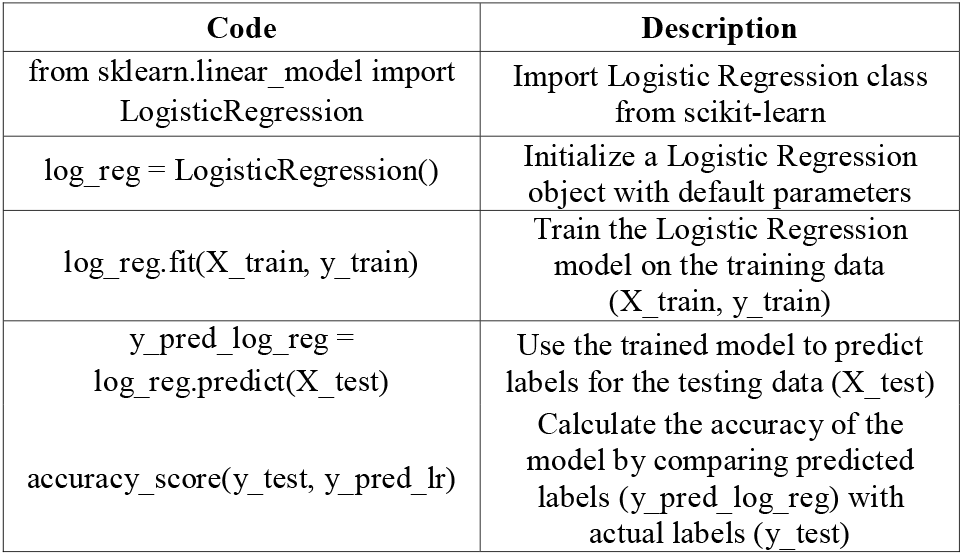
Logistic Regression Classifier Trained with Original Data.

| Code | Description |
| --- | --- |
| from sklearn.linear_model import<br>LogisticRegression | Import Logistic Regression class from scikit-learn |
| log_reg = LogisticRegression() | Initialize a Logistic Regression object with default parameters |
| log_reg.fit(X_train, y_train) | Train the Logistic Regression model on the training data (X_train, y_train) |
| y_pred_log_reg =<br>log_reg.predict(X_test) | Use the trained model to predict labels for the testing data (X_test) |
| accuracy_score(y_test, y_pred_lr) | Calculate the accuracy of the model by comparing predicted labels (y_pred_log_reg) with actual labels (y_test) |

**TABLE XII.** Logistic Regression Classifier Trained with Scaled Data.

| Code | Description |
| --- | --- |
| log_reg_sc =<br>LogisticRegression() | Initialize another Logistic Regression object for scaled data |
| log_reg_sc.fit(X_train_sc,<br>y_train) | Train the Logistic Regression model on scaled training data (X_train_sc, y_train) |
| y_pred_log_reg_sc =<br>log_reg_sc.predict(X_test_sc) | Use the trained model to predict labels for scaled testing data (X_test_sc) |
| accuracy_score(y_test,<br>y_pred_lr_sc) | Calculate the accuracy of the model on scaled data by comparing predicted labels (y_pred_log_reg_sc) with actual labels (y_test) |

#### c) K-Nearest Neighbor (KNN) Classifier

The KNN was trained with both the original and scaled data. Training the model with the original data is presented in Table 13, and training with scaled data is presented in Table 14.

**TABLE XIII.** K-Nearest Neighbor (KNN) Classifier Trained with Original Data.

| Code | Description |
| --- | --- |
| from sklearn.neighbors import<br>KNeighborsClassifier | Import K-Nearest Neighbors (KNN) Classifier from scikit-learn |
| knn = KNeighborsClassifier() | Initialize a KNN Classifier object with default parameters (k=5) |
| knn.fit(X_train, y_train) | Train the KNN model on the training data (X_train, y_train) |
| y_pred_knn = knn.predict(X_test) | Use the trained model to predict labels for the testing data (X_test) |
| accuracy_score(y_test, y_pred_knn) | Calculate the accuracy of the model by comparing predicted labels (y_pred_knn) with actual labels (y_test) |

**TABLE XIV.** K-Nearest Neighbor (KNN) Classifier Trained with Scaled Data.

| Code | Description |
| --- | --- |
| knn_sc = KNeighborsClassifier() | Initialize another KNN Classifier object for scaled data |
| knn_sc.fit(X_train_sc, y_train) | Train the KNN model on scaled training data (X_train_sc, y_train) |
| y_pred_knn_sc = knn_sc.predict(X_test_sc) | Use the trained model to predict labels for scaled testing data (X_test_sc) |
| accuracy_score(y_test, y_pred_knn_sc) | Calculate the accuracy of the model on scaled data by comparing predicted labels (y_pred_knn_sc) with actual labels (y_test) |

#### d) Naive Bayes Classifier

The Naive Bayes Classifier was trained with both the original and scaled data. Training the model with the original data is presented in Table 15, and training with scaled data is presented in Table 16.

**TABLE XV.**
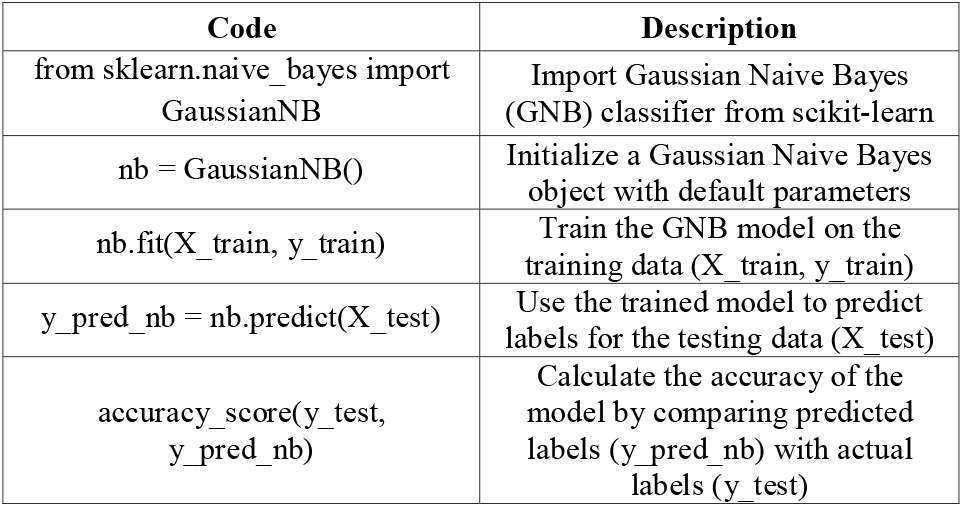
Naive Bayes Classifier Trained with Original Data.

**TABLE XVI.** Naive Bayes Classifier Trained with Scaled Data.

| Code | Description |
| --- | --- |
| nb_sc = GaussianNB() | Initialize another Gaussian Naive Bayes object for scaled data |
| nb_sc.fit(X_train_sc, y_train) | Train the GNB model on scaled training data (X_train_sc, y_train) |
| y_pred_nb_sc = nb_sc.predict(X_test_sc) | Use the trained model to predict labels for scaled testing data (X_test_sc) |
| accuracy_score(y_test, y_pred_nb_sc) | Calculate the accuracy of the model on scaled data by comparing predicted labels (y_pred_nb_sc) with actual labels (y_test) |

#### e) Decision Tree Classifier

The DTC was trained with both the original and scaled data. Training the model with the original data is presented in Table 17, and training with scaled data is presented in Table 18.

**TABLE XVII.** Decision Tree Classifier Trained with Original Data.

| Code | Description |
| --- | --- |
| from sklearn.tree import<br>DecisionTreeClassifier | Import Decision Tree Classifier from scikit-learn |
| dt = DecisionTreeClassifier() | Initialize a Decision Tree Classifier object with default parameters |
| dt.fit(X_train, y_train) | Train the Decision Tree model on the training data (X_train, y_train) |
| y_pred_dt = dt.predict(X_test) | Use the trained model to predict labels for the testing data (X_test) |
| accuracy_score(y_test, y_pred_dt) | Calculate the accuracy of the model by comparing predicted labels (y_pred_dt) with actual labels (y_test) |

**TABLE XVIII.**
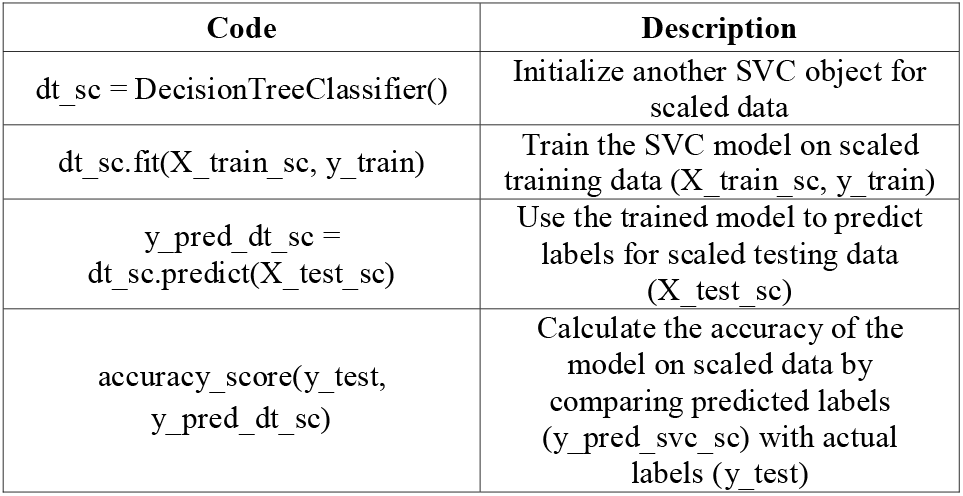
Decision Tree Classifier Trained with Scaled Data.

#### f) Random Forest Classifier

The RFC was trained with both the original and scaled data. Training the model with the original data is presented in Table 19, and training with scaled data is presented in Table 20.

**TABLE XIX.** Random Forest Classifier Trained with Original Data.

| Code | Description |
| --- | --- |
| from sklearn.ensemble import<br>RandomForestClassifier | Import Random Forest Classifier from scikit-learn |
| rf = RandomForestClassifier() | Initialize a Random Forest Classifier object with default parameters |
| rf.fit(X_train, y_train) | Train the Random Forest model on the training data (X_train, y_train) |
| y_pred_rf = rf.predict(X_test) | Use the trained model to predict labels for the testing data (X_test) |
| accuracy_score(y_test, y_pred_rf) | Calculate the accuracy of the model by comparing predicted labels (y_pred_rf) with actual labels (y_test) |

**TABLE XX.**
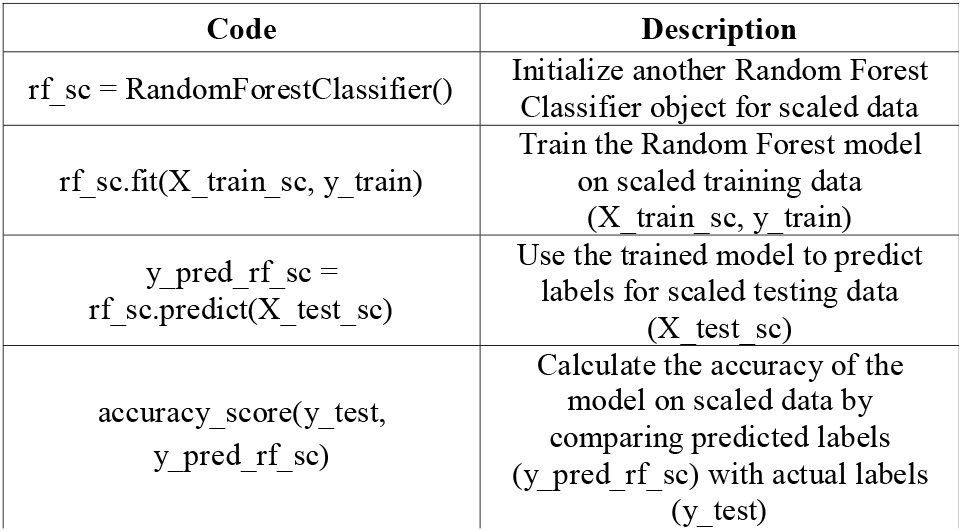
Random Forest Classifier Trained with Scaled Data.

| Code | Description |
| --- | --- |
| rf_sc = RandomForestClassifier() | Initialize another Random Forest Classifier object for scaled data |
| rf_sc.fit(X_train_sc, y_train) | Train the Random Forest model on scaled training data (X_train_sc, y_train) |
| y_pred_rf_sc = rf_sc.predict(X_test_sc) | Use the trained model to predict labels for scaled testing data (X_test_sc) |
| accuracy_score(y_test, y_pred_rf_sc) | Calculate the accuracy of the model on scaled data by comparing predicted labels (y_pred_rf_sc) with actual labels (y_test) |

### H. Result

In the study, several traditional machine learning algorithms were evaluated on a dataset comprising numerical features related to brain tumor detection. The performance of each algorithm was assessed both on the original data and after applying data scaling techniques to standardize the features. The results demonstrated significant improvements in model accuracy post-scaling.

The Support Vector Classifier (SVC) achieved a baseline accuracy of 78.61% on the original data. However, after scaling, the model’s performance surged to 97.74%, highlighting the crucial role of data scaling in boosting accuracy.

Logistic Regression Classifier demonstrated a notable accuracy of 90.96% on the original data. When applied to scaled data, the model’s performance further improved to 97.87%, indicating enhanced class separation and more effective feature utilization.

The K-Nearest Neighbors (KNN) Classifier exhibited an accuracy of 80.61% on the original data. However, its performance improved to 97.74% on scaled data, suggesting a positive impact from the scaling method employed.

Naive Bayes Classifier achieved a commendable accuracy of 95.35% on the original data. Although scaling had a minimal impact, the model’s accuracy edged up to 95.48%, indicating a slight benefit from standardized features.

The Decision Tree Classifier showcased an impressive accuracy of 98.14% on the original data, and this accuracy remained the same at 98.14% on scaled data, indicating no significant impact from scaling.

Lastly, the Random Forest Classifier achieved an outstanding accuracy of 98.27% on the original data, and this accuracy remained the same at 98.27% on scaled data, suggesting no significant impact from scaling.

The summarized results of all classifiers in Table 21:

**TABLE XXI.** Summarized result of all classifiers.

| Classifiers | Accuracy (Original Data) | Accuracy (Scaled Data) |
| --- | --- | --- |
| Support Vector Machine | 78.61% | 97.74% |
| Logistic Regression | 90.96% | 97.87% |
| K-Nearest Neighbors | 80.61% | 97.74% |
| Naive Bayes | 95.35% | 95.48% |
| Decision Tree | 98.14% | 98.14% |
| Random Forest | 98.27% | 98.27% |

### I. Discussion

In this study, we conducted a comprehensive evaluation of six machine learning classifiers: Support Vector Classifier (SVC), Logistic Regression, K-Nearest Neighbors (KNN), Naive Bayes, Decision Tree, and Random Forest on a brain tumor dataset, with a specific focus on the impact of data preprocessing on performance. Our results underscore the critical role of feature scaling in enhancing the performance of machine learning classifiers in medical diagnostics, highlighting Random Forest’s potential as a reliable tool for automated brain tumor detection.

#### a) Classifier Performance

While Random Forest achieved the highest accuracy of 98.27% on both original and scaled data, emerging as the top performer in both scenarios, this highlights the importance of feature scaling in unlocking the full potential of classifiers.

#### b) Implementations for Clinical Application

The consistent high performance of Random Forest, combined with its robustness to data distribution, has important implications for clinical applications. Integrating preprocessing steps can significantly enhance diagnostic accuracy, particularly for classifiers that rely heavily on data distribution. This finding suggests that Random Forest has the potential to become a reliable model for automated brain tumor detection, offering a valuable tool for clinicians in diagnosing and treating brain tumors, due to its consistently high performance on both original and scaled data.

#### c) Future Work

Future research could investigate the use of advanced deep learning architectures to improve tumor segmentation and detection. This includes exploring 3D CNNs for analyzing volumetric medical imaging data, and GANs for generating synthetic tumor data to augment real datasets. Additionally, multi-modal deep learning models could be developed to combine imaging data with clinical and genomic information. Transfer learning and attention mechanisms could also be examined to adapt pre-trained models to specific brain tumor types and improve detection accuracy. By evaluating these approaches, researchers can work towards enabling early intervention and personalized treatment strategies.

### J. Conclusion

This comprehensive study demonstrated the significance of data preprocessing in enhancing the performance of machine learning classifiers for brain tumor detection. The evaluation of six traditional machine learning algorithms - Support Vector Classifier (SVC), Logistic Regression, K-Nearest Neighbors (KNN), Naive Bayes, Decision Tree, and Random Forest - revealed that Random Forest emerged as the top performer, achieving the highest accuracy of 98.27% on both original and scaled data, showcasing its robustness and reliability. The findings suggest that Random Forest has the potential to become a reliable model for automated brain tumor detection, offering a valuable tool for clinicians. The consistent high performance of Random Forest, combined with its robustness to data distribution, has significant implications for clinical applications, suggesting its potential as a reliable model for automated brain tumor detection. Future research could explore advanced deep learning architectures, including 3D CNNs, GANs, and multi-modal models, to further improve tumor segmentation and detection accuracy. Additionally, investigating transfer learning and attention mechanisms could help adapt pre-trained models to specific brain tumor types, ultimately enabling early intervention and personalized treatment strategies.

## Supporting information

contains: supplemental file, brain_tumor_detection_model.ipynb, brain_tumor_detection_model.html, brain_tumor_dataset.csv

## Data Availability

https://www.kaggle.com/datasets/jakeshbohaju/brain-tumor

## Notes

### Competing Interest Statement

The authors have declared no competing interest.

### Clinical Protocols

https://github.com/1umairali/models/blob/main/brain_tumor_detection/brain_dataset.csv

https://www.kaggle.com/datasets/jakeshbohaju/brain-tumor

### Funding Statement

This study did not receive any funding

### Author Declarations

Dataset downloaded from: https://github.com/1umairali/models/tree/main/brain_tumor_detection https://www.kaggle.com/datasets/jakeshbohaju/brain-tumor

### Summary of Updates

I am pleased to resubmit my revised manuscript, which addresses the concerns and errors identified in the previous version. Specifically: - I apologize for the oversight, but the abstract was not updated in this revised version. I will make sure to revise it accordingly in the next submission. I have carefully reviewed the manuscript to ensure accuracy, completeness, and consistency. I believe these revisions strengthen the manuscript and provide a clearer presentation of my research.

