## Supplementary material for "Comparative Evaluation Of Machine Learning Classifiers For Brain Tumor Detection": contains: supplemental file, brain_tumor_detection_model.ipynb, brain_tumor_detection_model.html, brain_tumor_dataset.csv: brain_tumor_detection_model.html


### Brain Tumor Detection¶

**Model Developed by**   
Name: **Umair Ali**   
Contact: **+923480233673**  
GitHub:  **https://github.com/1umairali/models/tree/main/brain\_tumor\_detection**   
ORCID ID: **https://orcid.org/0009-0001-5156-4733**

In [1]:

```
import numpy as np  # for numeric calculation
import pandas as pd  # for data analysis and manupulation
import matplotlib.pyplot as plt  # for data visualization
import seaborn as sns  # for data visualization
```

In [2]:

```
# import dataframe
url = 'https://raw.githubusercontent.com/1umairali/models/main/brain_tumor_detection/brain_tumor_dataset.csv'
brain_dataframe = pd.read_csv(url)
brain_dataframe
```

Out[2]:

|  | Image | Class | Mean | Variance | Standard Deviation | Entropy | Skewness | Kurtosis | Contrast | Energy | ASM | Homogeneity | Dissimilarity | Correlation | Coarseness |
| --- | --- | --- | --- | --- | --- | --- | --- | --- | --- | --- | --- | --- | --- | --- | --- |
| 0 | Image1 | 0 | 6.535339 | 619.587845 | 24.891522 | 0.109059 | 4.276477 | 18.900575 | 98.613971 | 0.293314 | 0.086033 | 0.530941 | 4.473346 | 0.981939 | 7.458341e-155 |
| 1 | Image2 | 0 | 8.749969 | 805.957634 | 28.389393 | 0.266538 | 3.718116 | 14.464618 | 63.858816 | 0.475051 | 0.225674 | 0.651352 | 3.220072 | 0.988834 | 7.458341e-155 |
| 2 | Image3 | 1 | 7.341095 | 1143.808219 | 33.820234 | 0.001467 | 5.061750 | 26.479563 | 81.867206 | 0.031917 | 0.001019 | 0.268275 | 5.981800 | 0.978014 | 7.458341e-155 |
| 3 | Image4 | 1 | 5.958145 | 959.711985 | 30.979219 | 0.001477 | 5.677977 | 33.428845 | 151.229741 | 0.032024 | 0.001026 | 0.243851 | 7.700919 | 0.964189 | 7.458341e-155 |
| 4 | Image5 | 0 | 7.315231 | 729.540579 | 27.010009 | 0.146761 | 4.283221 | 19.079108 | 174.988756 | 0.343849 | 0.118232 | 0.501140 | 6.834689 | 0.972789 | 7.458341e-155 |
| ... | ... | ... | ... | ... | ... | ... | ... | ... | ... | ... | ... | ... | ... | ... | ... |
| 3757 | Image3758 | 0 | 21.234512 | 1208.850174 | 34.768523 | 0.063774 | 2.082079 | 4.647310 | 158.437600 | 0.220666 | 0.048693 | 0.487131 | 5.211739 | 0.950972 | 7.458341e-155 |
| 3758 | Image3759 | 0 | 20.435349 | 1227.151440 | 35.030721 | 0.066763 | 2.144625 | 4.882034 | 161.158675 | 0.225931 | 0.051045 | 0.502712 | 5.083126 | 0.952749 | 7.458341e-155 |
| 3759 | Image3760 | 0 | 18.011520 | 1151.582765 | 33.934978 | 0.068396 | 2.308349 | 5.579498 | 167.130118 | 0.228930 | 0.052409 | 0.492269 | 5.103700 | 0.952181 | 7.458341e-155 |
| 3760 | Image3761 | 0 | 13.330429 | 945.732779 | 30.752769 | 0.087872 | 2.732822 | 7.757570 | 223.812932 | 0.261527 | 0.068397 | 0.480064 | 6.439784 | 0.940898 | 7.458341e-155 |
| 3761 | Image3762 | 0 | 6.110138 | 480.884025 | 21.929068 | 0.118171 | 4.110669 | 17.538826 | 239.251388 | 0.306224 | 0.093773 | 0.494333 | 6.787329 | 0.938731 | 7.458341e-155 |

3762 rows × 15 columns

In [3]:

```
# Head (first six rows) of brain dataframe
brain_dataframe.head(6)
```

Out[3]:

|  | Image | Class | Mean | Variance | Standard Deviation | Entropy | Skewness | Kurtosis | Contrast | Energy | ASM | Homogeneity | Dissimilarity | Correlation | Coarseness |
| --- | --- | --- | --- | --- | --- | --- | --- | --- | --- | --- | --- | --- | --- | --- | --- |
| 0 | Image1 | 0 | 6.535339 | 619.587845 | 24.891522 | 0.109059 | 4.276477 | 18.900575 | 98.613971 | 0.293314 | 0.086033 | 0.530941 | 4.473346 | 0.981939 | 7.458341e-155 |
| 1 | Image2 | 0 | 8.749969 | 805.957634 | 28.389393 | 0.266538 | 3.718116 | 14.464618 | 63.858816 | 0.475051 | 0.225674 | 0.651352 | 3.220072 | 0.988834 | 7.458341e-155 |
| 2 | Image3 | 1 | 7.341095 | 1143.808219 | 33.820234 | 0.001467 | 5.061750 | 26.479563 | 81.867206 | 0.031917 | 0.001019 | 0.268275 | 5.981800 | 0.978014 | 7.458341e-155 |
| 3 | Image4 | 1 | 5.958145 | 959.711985 | 30.979219 | 0.001477 | 5.677977 | 33.428845 | 151.229741 | 0.032024 | 0.001026 | 0.243851 | 7.700919 | 0.964189 | 7.458341e-155 |
| 4 | Image5 | 0 | 7.315231 | 729.540579 | 27.010009 | 0.146761 | 4.283221 | 19.079108 | 174.988756 | 0.343849 | 0.118232 | 0.501140 | 6.834689 | 0.972789 | 7.458341e-155 |
| 5 | Image6 | 0 | 7.524109 | 607.395258 | 24.645390 | 0.214086 | 3.729886 | 14.471736 | 105.077882 | 0.421587 | 0.177736 | 0.598169 | 4.193146 | 0.976485 | 7.458341e-155 |

In [4]:

```
# Tail (last six rows) of brain dataframe
brain_dataframe.tail(6)
```

Out[4]:

|  | Image | Class | Mean | Variance | Standard Deviation | Entropy | Skewness | Kurtosis | Contrast | Energy | ASM | Homogeneity | Dissimilarity | Correlation | Coarseness |
| --- | --- | --- | --- | --- | --- | --- | --- | --- | --- | --- | --- | --- | --- | --- | --- |
| 3756 | Image3757 | 0 | 20.976822 | 1144.456066 | 33.829810 | 0.062252 | 2.106235 | 4.798339 | 166.395916 | 0.217934 | 0.047495 | 0.488449 | 5.193088 | 0.948808 | 7.458341e-155 |
| 3757 | Image3758 | 0 | 21.234512 | 1208.850174 | 34.768523 | 0.063774 | 2.082079 | 4.647310 | 158.437600 | 0.220666 | 0.048693 | 0.487131 | 5.211739 | 0.950972 | 7.458341e-155 |
| 3758 | Image3759 | 0 | 20.435349 | 1227.151440 | 35.030721 | 0.066763 | 2.144625 | 4.882034 | 161.158675 | 0.225931 | 0.051045 | 0.502712 | 5.083126 | 0.952749 | 7.458341e-155 |
| 3759 | Image3760 | 0 | 18.011520 | 1151.582765 | 33.934978 | 0.068396 | 2.308349 | 5.579498 | 167.130118 | 0.228930 | 0.052409 | 0.492269 | 5.103700 | 0.952181 | 7.458341e-155 |
| 3760 | Image3761 | 0 | 13.330429 | 945.732779 | 30.752769 | 0.087872 | 2.732822 | 7.757570 | 223.812932 | 0.261527 | 0.068397 | 0.480064 | 6.439784 | 0.940898 | 7.458341e-155 |
| 3761 | Image3762 | 0 | 6.110138 | 480.884025 | 21.929068 | 0.118171 | 4.110669 | 17.538826 | 239.251388 | 0.306224 | 0.093773 | 0.494333 | 6.787329 | 0.938731 | 7.458341e-155 |

In [5]:

```
# Information of brain dataframe
brain_dataframe.info()
```

```
<class 'pandas.core.frame.DataFrame'>
RangeIndex: 3762 entries, 0 to 3761
Data columns (total 15 columns):
 #   Column              Non-Null Count  Dtype  
---  ------              --------------  -----  
 0   Image               3762 non-null   object 
 1   Class               3762 non-null   int64  
 2   Mean                3762 non-null   float64
 3   Variance            3762 non-null   float64
 4   Standard Deviation  3762 non-null   float64
 5   Entropy             3762 non-null   float64
 6   Skewness            3762 non-null   float64
 7   Kurtosis            3762 non-null   float64
 8   Contrast            3762 non-null   float64
 9   Energy              3762 non-null   float64
 10  ASM                 3762 non-null   float64
 11  Homogeneity         3762 non-null   float64
 12  Dissimilarity       3762 non-null   float64
 13  Correlation         3762 non-null   float64
 14  Coarseness          3762 non-null   float64
dtypes: float64(13), int64(1), object(1)
memory usage: 441.0+ KB
```

In [6]:

```
# show image columns
print(brain_dataframe['Image'])
```

```
0          Image1
1          Image2
2          Image3
3          Image4
4          Image5
          ...    
3757    Image3758
3758    Image3759
3759    Image3760
3760    Image3761
3761    Image3762
Name: Image, Length: 3762, dtype: object
```

image column is object dtype. contains only images name. if we drop that column it will not impact dataframe or result

In [7]:

```
# drop Image column.
brain_df2 = brain_dataframe.drop(['Image'], axis = 1)
brain_df2
```

Out[7]:

|  | Class | Mean | Variance | Standard Deviation | Entropy | Skewness | Kurtosis | Contrast | Energy | ASM | Homogeneity | Dissimilarity | Correlation | Coarseness |
| --- | --- | --- | --- | --- | --- | --- | --- | --- | --- | --- | --- | --- | --- | --- |
| 0 | 0 | 6.535339 | 619.587845 | 24.891522 | 0.109059 | 4.276477 | 18.900575 | 98.613971 | 0.293314 | 0.086033 | 0.530941 | 4.473346 | 0.981939 | 7.458341e-155 |
| 1 | 0 | 8.749969 | 805.957634 | 28.389393 | 0.266538 | 3.718116 | 14.464618 | 63.858816 | 0.475051 | 0.225674 | 0.651352 | 3.220072 | 0.988834 | 7.458341e-155 |
| 2 | 1 | 7.341095 | 1143.808219 | 33.820234 | 0.001467 | 5.061750 | 26.479563 | 81.867206 | 0.031917 | 0.001019 | 0.268275 | 5.981800 | 0.978014 | 7.458341e-155 |
| 3 | 1 | 5.958145 | 959.711985 | 30.979219 | 0.001477 | 5.677977 | 33.428845 | 151.229741 | 0.032024 | 0.001026 | 0.243851 | 7.700919 | 0.964189 | 7.458341e-155 |
| 4 | 0 | 7.315231 | 729.540579 | 27.010009 | 0.146761 | 4.283221 | 19.079108 | 174.988756 | 0.343849 | 0.118232 | 0.501140 | 6.834689 | 0.972789 | 7.458341e-155 |
| ... | ... | ... | ... | ... | ... | ... | ... | ... | ... | ... | ... | ... | ... | ... |
| 3757 | 0 | 21.234512 | 1208.850174 | 34.768523 | 0.063774 | 2.082079 | 4.647310 | 158.437600 | 0.220666 | 0.048693 | 0.487131 | 5.211739 | 0.950972 | 7.458341e-155 |
| 3758 | 0 | 20.435349 | 1227.151440 | 35.030721 | 0.066763 | 2.144625 | 4.882034 | 161.158675 | 0.225931 | 0.051045 | 0.502712 | 5.083126 | 0.952749 | 7.458341e-155 |
| 3759 | 0 | 18.011520 | 1151.582765 | 33.934978 | 0.068396 | 2.308349 | 5.579498 | 167.130118 | 0.228930 | 0.052409 | 0.492269 | 5.103700 | 0.952181 | 7.458341e-155 |
| 3760 | 0 | 13.330429 | 945.732779 | 30.752769 | 0.087872 | 2.732822 | 7.757570 | 223.812932 | 0.261527 | 0.068397 | 0.480064 | 6.439784 | 0.940898 | 7.458341e-155 |
| 3761 | 0 | 6.110138 | 480.884025 | 21.929068 | 0.118171 | 4.110669 | 17.538826 | 239.251388 | 0.306224 | 0.093773 | 0.494333 | 6.787329 | 0.938731 | 7.458341e-155 |

3762 rows × 14 columns

In [8]:

```
# Numerical distribution of data
brain_df2.describe()
```

Out[8]:

|  | Class | Mean | Variance | Standard Deviation | Entropy | Skewness | Kurtosis | Contrast | Energy | ASM | Homogeneity | Dissimilarity | Correlation | Coarseness |
| --- | --- | --- | --- | --- | --- | --- | --- | --- | --- | --- | --- | --- | --- | --- |
| count | 3762.000000 | 3762.000000 | 3762.000000 | 3762.000000 | 3762.000000 | 3762.000000 | 3762.000000 | 3762.000000 | 3762.000000 | 3762.000000 | 3762.000000 | 3762.000000 | 3762.000000 | 3.762000e+03 |
| mean | 0.447368 | 9.488890 | 711.101063 | 25.182271 | 0.073603 | 4.102727 | 24.389071 | 127.961459 | 0.204705 | 0.058632 | 0.479252 | 4.698498 | 0.955767 | 7.458341e-155 |
| std | 0.497288 | 5.728022 | 467.466896 | 8.773526 | 0.070269 | 2.560940 | 56.434747 | 109.499601 | 0.129352 | 0.058300 | 0.127929 | 1.850173 | 0.026157 | 0.000000e+00 |
| min | 0.000000 | 0.078659 | 3.145628 | 1.773592 | 0.000882 | 1.886014 | 3.942402 | 3.194733 | 0.024731 | 0.000612 | 0.105490 | 0.681121 | 0.549426 | 7.458341e-155 |
| 25% | 0.000000 | 4.982395 | 363.225459 | 19.058475 | 0.006856 | 2.620203 | 7.252852 | 72.125208 | 0.069617 | 0.004847 | 0.364973 | 3.412363 | 0.947138 | 7.458341e-155 |
| 50% | 0.000000 | 8.477531 | 622.580417 | 24.951560 | 0.066628 | 3.422210 | 12.359088 | 106.737418 | 0.225496 | 0.050849 | 0.512551 | 4.482404 | 0.961610 | 7.458341e-155 |
| 75% | 1.000000 | 13.212723 | 966.954319 | 31.095889 | 0.113284 | 4.651737 | 22.640304 | 161.059006 | 0.298901 | 0.089342 | 0.575557 | 5.723821 | 0.971355 | 7.458341e-155 |
| max | 1.000000 | 33.239975 | 2910.581879 | 53.949809 | 0.394539 | 36.931294 | 1371.640060 | 3382.574163 | 0.589682 | 0.347725 | 0.810921 | 27.827751 | 0.989972 | 7.458341e-155 |

In [9]:

```
# check sum of null values in each columns
brain_df2.isnull().sum()
```

Out[9]:

```
Class                 0
Mean                  0
Variance              0
Standard Deviation    0
Entropy               0
Skewness              0
Kurtosis              0
Contrast              0
Energy                0
ASM                   0
Homogeneity           0
Dissimilarity         0
Correlation           0
Coarseness            0
dtype: int64
```

### Data Visualization¶

In [10]:

```
# Pairplot of brain dataframe
sns.pairplot(brain_df2, hue = 'Class')
```

```
C:\Users\Umair Ali\anaconda3\Lib\site-packages\seaborn\_oldcore.py:1119: FutureWarning: use_inf_as_na option is deprecated and will be removed in a future version. Convert inf values to NaN before operating instead.
  with pd.option_context('mode.use_inf_as_na', True):
C:\Users\Umair Ali\anaconda3\Lib\site-packages\seaborn\_oldcore.py:1119: FutureWarning: use_inf_as_na option is deprecated and will be removed in a future version. Convert inf values to NaN before operating instead.
  with pd.option_context('mode.use_inf_as_na', True):
C:\Users\Umair Ali\anaconda3\Lib\site-packages\seaborn\_oldcore.py:1119: FutureWarning: use_inf_as_na option is deprecated and will be removed in a future version. Convert inf values to NaN before operating instead.
  with pd.option_context('mode.use_inf_as_na', True):
C:\Users\Umair Ali\anaconda3\Lib\site-packages\seaborn\_oldcore.py:1119: FutureWarning: use_inf_as_na option is deprecated and will be removed in a future version. Convert inf values to NaN before operating instead.
  with pd.option_context('mode.use_inf_as_na', True):
C:\Users\Umair Ali\anaconda3\Lib\site-packages\seaborn\_oldcore.py:1119: FutureWarning: use_inf_as_na option is deprecated and will be removed in a future version. Convert inf values to NaN before operating instead.
  with pd.option_context('mode.use_inf_as_na', True):
C:\Users\Umair Ali\anaconda3\Lib\site-packages\seaborn\_oldcore.py:1119: FutureWarning: use_inf_as_na option is deprecated and will be removed in a future version. Convert inf values to NaN before operating instead.
  with pd.option_context('mode.use_inf_as_na', True):
C:\Users\Umair Ali\anaconda3\Lib\site-packages\seaborn\_oldcore.py:1119: FutureWarning: use_inf_as_na option is deprecated and will be removed in a future version. Convert inf values to NaN before operating instead.
  with pd.option_context('mode.use_inf_as_na', True):
C:\Users\Umair Ali\anaconda3\Lib\site-packages\seaborn\_oldcore.py:1119: FutureWarning: use_inf_as_na option is deprecated and will be removed in a future version. Convert inf values to NaN before operating instead.
  with pd.option_context('mode.use_inf_as_na', True):
C:\Users\Umair Ali\anaconda3\Lib\site-packages\seaborn\_oldcore.py:1119: FutureWarning: use_inf_as_na option is deprecated and will be removed in a future version. Convert inf values to NaN before operating instead.
  with pd.option_context('mode.use_inf_as_na', True):
C:\Users\Umair Ali\anaconda3\Lib\site-packages\seaborn\_oldcore.py:1119: FutureWarning: use_inf_as_na option is deprecated and will be removed in a future version. Convert inf values to NaN before operating instead.
  with pd.option_context('mode.use_inf_as_na', True):
C:\Users\Umair Ali\anaconda3\Lib\site-packages\seaborn\_oldcore.py:1119: FutureWarning: use_inf_as_na option is deprecated and will be removed in a future version. Convert inf values to NaN before operating instead.
  with pd.option_context('mode.use_inf_as_na', True):
C:\Users\Umair Ali\anaconda3\Lib\site-packages\seaborn\_oldcore.py:1119: FutureWarning: use_inf_as_na option is deprecated and will be removed in a future version. Convert inf values to NaN before operating instead.
  with pd.option_context('mode.use_inf_as_na', True):
C:\Users\Umair Ali\anaconda3\Lib\site-packages\seaborn\_oldcore.py:1119: FutureWarning: use_inf_as_na option is deprecated and will be removed in a future version. Convert inf values to NaN before operating instead.
  with pd.option_context('mode.use_inf_as_na', True):
```

Out[10]:

```
<seaborn.axisgrid.PairGrid at 0x29d0ea37c50>
```

In [11]:

```
# Count the class columns
# 0 = Non tumor / no cancer
# 1 = Tumor / has cancer
sns.countplot(x=brain_df2["Class"])
```

Out[11]:

```
<Axes: xlabel='Class', ylabel='count'>
```

### Heatmap¶

In [12]:

```
# heatmap of DataFrame
plt.figure(figsize=(16,9))
sns.heatmap(brain_df2)
```

Out[12]:

```
<Axes: >
```

### Heatmap of a correlation Matrix¶

In [13]:

```
# correlation matrix
brain_df2.corr()
```

Out[13]:

|  | Class | Mean | Variance | Standard Deviation | Entropy | Skewness | Kurtosis | Contrast | Energy | ASM | Homogeneity | Dissimilarity | Correlation | Coarseness |
| --- | --- | --- | --- | --- | --- | --- | --- | --- | --- | --- | --- | --- | --- | --- |
| Class | 1.000000 | -0.095729 | 0.308818 | 0.285568 | -0.778180 | 0.402644 | 0.239844 | 0.212643 | -0.862413 | -0.758255 | -0.847529 | 0.556319 | -0.108601 | NaN |
| Mean | -0.095729 | 1.000000 | 0.783027 | 0.790984 | -0.099729 | -0.601593 | -0.358163 | -0.050974 | -0.014863 | -0.109393 | 0.095556 | -0.113864 | 0.293693 | NaN |
| Variance | 0.308818 | 0.783027 | 1.000000 | 0.975699 | -0.344432 | -0.347399 | -0.248312 | 0.135494 | -0.335470 | -0.341061 | -0.290527 | 0.235487 | 0.288037 | NaN |
| Standard Deviation | 0.285568 | 0.790984 | 0.975699 | 1.000000 | -0.345127 | -0.425428 | -0.329798 | 0.117981 | -0.331103 | -0.342530 | -0.288801 | 0.224773 | 0.354161 | NaN |
| Entropy | -0.778180 | -0.099729 | -0.344432 | -0.345127 | 1.000000 | -0.222222 | -0.140125 | -0.140769 | 0.971260 | 0.999213 | 0.852019 | -0.502363 | 0.122080 | NaN |
| Skewness | 0.402644 | -0.601593 | -0.347399 | -0.425428 | -0.222222 | 1.000000 | 0.899713 | 0.349856 | -0.295413 | -0.209289 | -0.470054 | 0.511931 | -0.570919 | NaN |
| Kurtosis | 0.239844 | -0.358163 | -0.248312 | -0.329798 | -0.140125 | 0.899713 | 1.000000 | 0.296664 | -0.172454 | -0.133741 | -0.307314 | 0.375939 | -0.589211 | NaN |
| Contrast | 0.212643 | -0.050974 | 0.135494 | 0.117981 | -0.140769 | 0.349856 | 0.296664 | 1.000000 | -0.130708 | -0.139276 | -0.270119 | 0.761497 | -0.427443 | NaN |
| Energy | -0.862413 | -0.014863 | -0.335470 | -0.331103 | 0.971260 | -0.295413 | -0.172454 | -0.130708 | 1.000000 | 0.961628 | 0.915988 | -0.545774 | 0.123680 | NaN |
| ASM | -0.758255 | -0.109393 | -0.341061 | -0.342530 | 0.999213 | -0.209289 | -0.133741 | -0.139276 | 0.961628 | 1.000000 | 0.837139 | -0.491813 | 0.121054 | NaN |
| Homogeneity | -0.847529 | 0.095556 | -0.290527 | -0.288801 | 0.852019 | -0.470054 | -0.307314 | -0.270119 | 0.915988 | 0.837139 | 1.000000 | -0.746675 | 0.198639 | NaN |
| Dissimilarity | 0.556319 | -0.113864 | 0.235487 | 0.224773 | -0.502363 | 0.511931 | 0.375939 | 0.761497 | -0.545774 | -0.491813 | -0.746675 | 1.000000 | -0.393013 | NaN |
| Correlation | -0.108601 | 0.293693 | 0.288037 | 0.354161 | 0.122080 | -0.570919 | -0.589211 | -0.427443 | 0.123680 | 0.121054 | 0.198639 | -0.393013 | 1.000000 | NaN |
| Coarseness | NaN | NaN | NaN | NaN | NaN | NaN | NaN | NaN | NaN | NaN | NaN | NaN | NaN | NaN |

we checked in information 'Coarseness' column is float datatype. contains 0 values which is not empty.......
but in corelation matrix it contain NaN values......
we have to remove that column. it will not impact the result......
lets confirm in Heatmap and Barplot of Correlation Matrix

In [14]:

```
# Heatmap of Correlation matrix of Brain DataFrame
plt.figure(figsize=(10,8))
sns.heatmap(brain_df2.corr(), annot = True, cmap ='coolwarm', linewidths=2)
```

```
C:\Users\Umair Ali\anaconda3\Lib\site-packages\seaborn\matrix.py:260: FutureWarning: Format strings passed to MaskedConstant are ignored, but in future may error or produce different behavior
  annotation = ("{:" + self.fmt + "}").format(val)
```

Out[14]:

```
<Axes: >
```

In [15]:

```
# drop coarseness column from brain_df2
brain_df3 = brain_df2.drop(['Coarseness'], axis=1)
```

### Split Dataframe in Train and Test¶

In [16]:

```
# drop dependent (Class) column, it will assign to y
X = brain_df3.drop(['Class'], axis = 1)
X.head(6)
```

Out[16]:

|  | Mean | Variance | Standard Deviation | Entropy | Skewness | Kurtosis | Contrast | Energy | ASM | Homogeneity | Dissimilarity | Correlation |
| --- | --- | --- | --- | --- | --- | --- | --- | --- | --- | --- | --- | --- |
| 0 | 6.535339 | 619.587845 | 24.891522 | 0.109059 | 4.276477 | 18.900575 | 98.613971 | 0.293314 | 0.086033 | 0.530941 | 4.473346 | 0.981939 |
| 1 | 8.749969 | 805.957634 | 28.389393 | 0.266538 | 3.718116 | 14.464618 | 63.858816 | 0.475051 | 0.225674 | 0.651352 | 3.220072 | 0.988834 |
| 2 | 7.341095 | 1143.808219 | 33.820234 | 0.001467 | 5.061750 | 26.479563 | 81.867206 | 0.031917 | 0.001019 | 0.268275 | 5.981800 | 0.978014 |
| 3 | 5.958145 | 959.711985 | 30.979219 | 0.001477 | 5.677977 | 33.428845 | 151.229741 | 0.032024 | 0.001026 | 0.243851 | 7.700919 | 0.964189 |
| 4 | 7.315231 | 729.540579 | 27.010009 | 0.146761 | 4.283221 | 19.079108 | 174.988756 | 0.343849 | 0.118232 | 0.501140 | 6.834689 | 0.972789 |
| 5 | 7.524109 | 607.395258 | 24.645390 | 0.214086 | 3.729886 | 14.471736 | 105.077882 | 0.421587 | 0.177736 | 0.598169 | 4.193146 | 0.976485 |

In [17]:

```
# output variable
# assign Class column to y
y = brain_df3['Class']
y.head(6)
```

Out[17]:

```
0    0
1    0
2    1
3    1
4    0
5    0
Name: Class, dtype: int64
```

In [18]:

```
# split dataset into train and test
from sklearn.model_selection import train_test_split
X_train, X_test, y_train, y_test = train_test_split(X, y, test_size = 0.2, random_state= 5)
```

In [19]:

```
X_train
```

Out[19]:

|  | Mean | Variance | Standard Deviation | Entropy | Skewness | Kurtosis | Contrast | Energy | ASM | Homogeneity | Dissimilarity | Correlation |
| --- | --- | --- | --- | --- | --- | --- | --- | --- | --- | --- | --- | --- |
| 2021 | 7.885529 | 681.803866 | 26.111374 | 0.133134 | 3.806777 | 14.939540 | 277.900762 | 0.326207 | 0.106411 | 0.546301 | 6.445886 | 0.944215 |
| 1286 | 6.639282 | 207.782336 | 14.414657 | 0.102582 | 2.607407 | 7.151179 | 33.038741 | 0.282621 | 0.079875 | 0.638838 | 2.064030 | 0.959070 |
| 1106 | 3.186020 | 314.159477 | 17.724544 | 0.179837 | 6.354288 | 43.889536 | 165.700313 | 0.383583 | 0.147136 | 0.569095 | 4.329943 | 0.969997 |
| 3688 | 7.752167 | 850.780349 | 29.168139 | 0.000954 | 4.667157 | 23.797444 | 162.713111 | 0.025726 | 0.000662 | 0.255565 | 7.630112 | 0.966914 |
| 1781 | 15.907272 | 859.296845 | 29.313765 | 0.093459 | 2.257804 | 5.399814 | 111.219504 | 0.269897 | 0.072845 | 0.543017 | 3.945097 | 0.960212 |
| ... | ... | ... | ... | ... | ... | ... | ... | ... | ... | ... | ... | ... |
| 3190 | 19.755035 | 1511.266702 | 38.875014 | 0.098117 | 2.359856 | 5.823204 | 155.330111 | 0.276962 | 0.076708 | 0.564342 | 4.108485 | 0.970138 |
| 3046 | 17.384506 | 1151.899226 | 33.939641 | 0.007915 | 2.516995 | 6.645917 | 119.718675 | 0.075044 | 0.005632 | 0.381815 | 5.310392 | 0.944067 |
| 1725 | 9.873734 | 647.095405 | 25.438070 | 0.203971 | 2.936003 | 8.936402 | 116.379595 | 0.410201 | 0.168265 | 0.653499 | 3.168139 | 0.965933 |
| 2254 | 4.993439 | 837.608629 | 28.941469 | 0.001184 | 6.201616 | 39.682251 | 98.674332 | 0.028656 | 0.000821 | 0.218662 | 6.251872 | 0.960241 |
| 2915 | 14.946762 | 1063.536983 | 32.611915 | 0.144926 | 2.589608 | 7.070669 | 140.031810 | 0.341421 | 0.116569 | 0.579450 | 4.088308 | 0.968672 |

3009 rows × 12 columns

In [20]:

```
X_test
```

Out[20]:

|  | Mean | Variance | Standard Deviation | Entropy | Skewness | Kurtosis | Contrast | Energy | ASM | Homogeneity | Dissimilarity | Correlation |
| --- | --- | --- | --- | --- | --- | --- | --- | --- | --- | --- | --- | --- |
| 1829 | 14.244751 | 677.906461 | 26.036637 | 0.064730 | 2.245463 | 5.280704 | 68.194040 | 0.221802 | 0.049196 | 0.573020 | 3.171980 | 0.965574 |
| 142 | 6.439590 | 711.402252 | 26.672125 | 0.003683 | 4.945783 | 26.258025 | 191.833333 | 0.050998 | 0.002601 | 0.252045 | 8.161609 | 0.962899 |
| 2934 | 18.273666 | 960.554970 | 30.992821 | 0.053945 | 2.113554 | 4.673078 | 135.033939 | 0.201307 | 0.040524 | 0.550705 | 3.961433 | 0.943238 |
| 1648 | 8.556091 | 557.147920 | 23.603981 | 0.187408 | 3.215624 | 10.791892 | 87.087468 | 0.392090 | 0.153735 | 0.626582 | 3.201900 | 0.973064 |
| 1178 | 6.661285 | 761.907895 | 27.602679 | 0.004002 | 4.637791 | 22.559937 | 81.489763 | 0.052887 | 0.002797 | 0.370518 | 4.495690 | 0.969339 |
| ... | ... | ... | ... | ... | ... | ... | ... | ... | ... | ... | ... | ... |
| 3044 | 2.566971 | 254.252713 | 15.945304 | 0.001956 | 6.762827 | 47.224837 | 43.256424 | 0.036846 | 0.001358 | 0.317558 | 4.147622 | 0.983775 |
| 2743 | 11.161804 | 736.299788 | 27.134845 | 0.071535 | 2.928248 | 8.891376 | 147.695042 | 0.234516 | 0.054998 | 0.510246 | 5.120198 | 0.954483 |
| 3413 | 7.838409 | 300.574737 | 17.337091 | 0.134217 | 2.797981 | 8.655661 | 51.790533 | 0.327457 | 0.107228 | 0.616866 | 2.620355 | 0.960541 |
| 1619 | 4.232285 | 153.173766 | 12.376339 | 0.095976 | 3.288243 | 11.029013 | 35.134788 | 0.271702 | 0.073822 | 0.639419 | 2.224797 | 0.956256 |
| 3220 | 8.853958 | 573.067789 | 23.938834 | 0.006357 | 3.228923 | 10.874134 | 90.170569 | 0.067020 | 0.004492 | 0.391093 | 4.633593 | 0.939068 |

753 rows × 12 columns

In [21]:

```
y_train
```

Out[21]:

```
2021    0
1286    0
1106    1
3688    1
1781    0
       ..
3190    0
3046    1
1725    0
2254    1
2915    0
Name: Class, Length: 3009, dtype: int64
```

In [22]:

```
y_test
```

Out[22]:

```
1829    0
142     1
2934    0
1648    0
1178    1
       ..
3044    1
2743    0
3413    0
1619    0
3220    1
Name: Class, Length: 753, dtype: int64
```

### Feature Scaling¶

In [23]:

```
from sklearn.preprocessing import StandardScaler
sc = StandardScaler()
X_train_sc = sc.fit_transform(X_train)
X_test_sc = sc.transform(X_test)
```

### ML Model Building¶

In [24]:

```
from sklearn.metrics import confusion_matrix, classification_report, accuracy_score
```

### Support Vector Classifier¶

###### Train with original data¶

In [48]:

```
from sklearn.svm import SVC
svc_classifier = SVC()
svc_classifier.fit(X_train, y_train)
y_pred_svc = svc_classifier.predict(X_test)
accuracy_score(y_test, y_pred_svc)
```

Out[48]:

```
0.7861885790172642
```

###### Train with scaled data¶

In [49]:

```
svc_classifier2 = SVC()
svc_classifier2.fit(X_train_sc, y_train)
y_pred_svc_sc = svc_classifier2.predict(X_test_sc)
accuracy_score(y_test, y_pred_svc_sc)
```

Out[49]:

```
0.9774236387782205
```

### Logistic Regression¶

###### Train with original data¶

In [27]:

```
from sklearn.linear_model import LogisticRegression
lr_classifier = LogisticRegression(random_state=51, penalty = 'l2')
lr_classifier.fit(X_train, y_train)
y_pred_lr = lr_classifier.predict(X_test)
accuracy_score(y_test, y_pred_lr)
```

```
C:\Users\Umair Ali\anaconda3\Lib\site-packages\sklearn\linear_model\_logistic.py:458: ConvergenceWarning: lbfgs failed to converge (status=1):
STOP: TOTAL NO. of ITERATIONS REACHED LIMIT.

Increase the number of iterations (max_iter) or scale the data as shown in:
    https://scikit-learn.org/stable/modules/preprocessing.html
Please also refer to the documentation for alternative solver options:
    https://scikit-learn.org/stable/modules/linear_model.html#logistic-regression
  n_iter_i = _check_optimize_result(
```

Out[27]:

```
0.9096945551128818
```

###### Train with scaled data¶

In [50]:

```
lr_classifier2 = LogisticRegression(random_state=51, penalty = 'l2')
lr_classifier2.fit(X_train_sc, y_train)
y_pred_lr_sc = lr_classifier.predict(X_test_sc)
accuracy_score(y_test, y_pred_lr_sc)
```

```
C:\Users\Umair Ali\anaconda3\Lib\site-packages\sklearn\base.py:439: UserWarning: X does not have valid feature names, but LogisticRegression was fitted with feature names
  warnings.warn(
```

Out[50]:

```
0.9588313413014609
```

### KNN - K-Nearesr Neighbor Classifier¶

###### Train with original data¶

In [29]:

```
from sklearn.neighbors import KNeighborsClassifier
knn_classifier = KNeighborsClassifier(n_neighbors = 5, metric = 'minkowski', p = 2)
knn_classifier.fit(X_train, y_train)
y_pred_knn = knn_classifier.predict(X_test)
accuracy_score(y_test, y_pred_knn)
```

Out[29]:

```
0.8061088977423638
```

###### Train with scaled data¶

In [30]:

```
knn_classifier2 = KNeighborsClassifier(n_neighbors = 5, metric = 'minkowski', p = 2)
knn_classifier2.fit(X_train_sc, y_train)
y_pred_knn_sc = knn_classifier.predict(X_test_sc)
accuracy_score(y_test, y_pred_knn_sc)
```

```
C:\Users\Umair Ali\anaconda3\Lib\site-packages\sklearn\base.py:439: UserWarning: X does not have valid feature names, but KNeighborsClassifier was fitted with feature names
  warnings.warn(
```

Out[30]:

```
0.545816733067729
```

### Naive Bayes Classifier¶

###### Train with original data¶

In [45]:

```
from sklearn.naive_bayes import GaussianNB
nb_classifier = GaussianNB()
nb_classifier.fit(X_train, y_train)
y_pred_nb = nb_classifier.predict(X_test)
accuracy_score(y_test, y_pred_nb)
```

Out[45]:

```
0.953519256308101
```

###### Train with scaled data¶

In [46]:

```
nb_classifier2 = GaussianNB()
nb_classifier2.fit(X_train_sc, y_train)
y_pred_nb_sc = nb_classifier2.predict(X_test_sc)
accuracy_score(y_test, y_pred_nb_sc)
```

Out[46]:

```
0.9548472775564409
```

### Decision Tree Classifier¶

###### Train with original data¶

In [33]:

```
from sklearn.tree import DecisionTreeClassifier
dt_classifier = DecisionTreeClassifier(criterion = 'entropy', random_state = 51)
dt_classifier.fit(X_train, y_train)
y_pred_dt = dt_classifier.predict(X_test)
accuracy_score(y_test, y_pred_dt)
```

Out[33]:

```
0.9814077025232404
```

###### Train with scaled data¶

In [34]:

```
dt_classifier2 = DecisionTreeClassifier(criterion = 'entropy', random_state = 51)
dt_classifier2.fit(X_train_sc, y_train)
y_pred_dt_sc = dt_classifier.predict(X_test_sc)
accuracy_score(y_test, y_pred_dt_sc)
```

```
C:\Users\Umair Ali\anaconda3\Lib\site-packages\sklearn\base.py:439: UserWarning: X does not have valid feature names, but DecisionTreeClassifier was fitted with feature names
  warnings.warn(
```

Out[34]:

```
0.9030544488711819
```

### Random Forest Classifier¶

###### Train with original data¶

In [35]:

```
from sklearn.ensemble import RandomForestClassifier
rf_classifier = RandomForestClassifier(n_estimators = 20, criterion = 'entropy', random_state = 51)
rf_classifier.fit(X_train, y_train)
y_pred_rf = rf_classifier.predict(X_test)
accuracy_score(y_test, y_pred_rf)
```

Out[35]:

```
0.9827357237715804
```

###### Train with scaled data¶

In [36]:

```
rf_classifier2 = RandomForestClassifier(n_estimators = 20, criterion = 'entropy', random_state = 51)
rf_classifier2.fit(X_train_sc, y_train)
y_pred_rf_sc = rf_classifier.predict(X_test_sc)
accuracy_score(y_test, y_pred_rf_sc)
```

```
C:\Users\Umair Ali\anaconda3\Lib\site-packages\sklearn\base.py:439: UserWarning: X does not have valid feature names, but RandomForestClassifier was fitted with feature names
  warnings.warn(
```

Out[36]:

```
0.9389110225763613
```

### Confusion Matrix¶

In [37]:

```
cm = confusion_matrix(y_test, y_pred_svc_sc)
plt.title('Heatmap of Confusion Matrix', fontsize = 15)
sns.heatmap(cm, annot = True)
plt.show()
```

### Classification Report Of model¶

In [38]:

```
print(classification_report(y_test, y_pred_svc_sc))
```

```
              precision    recall  f1-score   support

           0       0.97      0.99      0.98       411
           1       0.99      0.96      0.97       342

    accuracy                           0.98       753
   macro avg       0.98      0.98      0.98       753
weighted avg       0.98      0.98      0.98       753
```

### Cross-validation of the ML model¶

In [39]:

```
# Cross validation
from sklearn.model_selection import cross_val_score
cross_validation = cross_val_score(estimator = svc_classifier2, X = X_train_sc,y = y_train, cv = 10)
print("Cross validation accuracy of SVC model = ", cross_validation)
print("\nCross validation mean accuracy of SVC model = ", cross_validation.mean())
```

```
Cross validation accuracy of SVC model =  [0.98671096 0.9833887  0.9833887  0.97342193 0.98671096 0.9833887
 0.9833887  0.9833887  0.99335548 0.99      ]

Cross validation mean accuracy of SVC model =  0.9847142857142858
```

### Test Model¶

In [40]:

```
# we have one patient data in list
patient1 = [17.99,10.38,122.8,1001.0,0.1184,0.2776,0.3001,
            0.1471,0.2419,0.07871,1.095,0.9053]
```

In [41]:

```
# convert list into array and scale
patient1_sc = sc.transform(np.array([patient1]))
patient1_sc
```

```
C:\Users\Umair Ali\anaconda3\Lib\site-packages\sklearn\base.py:439: UserWarning: X does not have valid feature names, but StandardScaler was fitted with feature names
  warnings.warn(
```

Out[41]:

```
array([[ 1.49256979e+00, -1.49381108e+00,  1.11029758e+01,
         1.40960224e+04, -1.54791906e+00, -4.16334494e-01,
        -1.13805522e+00, -4.48064734e-01,  3.09915646e+00,
        -3.10934480e+00, -1.93429813e+00, -1.84574993e+00]])
```

In [42]:

```
# predict patient1_sc scale data
# zero mean malignant patient has cancer
predict = svc_classifier2.predict(patient1_sc)
predict
```

Out[42]:

```
array([1], dtype=int64)
```

In [43]:

```
# write if else statement to print result in clear format
if predict[0] == 0:
    print ('Patient has *** No *** Tumor / Cancer')
else:
    print ('Patient *** has *** Tumor / Cancer')
```

```
Patient *** has *** Tumor / Cancer
```

In [44]:

```
# confusion matrix
print('Confusion matrix of SVC model: \n',confusion_matrix(y_test, y_pred_svc_sc),'\n')

# show the accuracy
print('Accuracy of SVC model = ',accuracy_score(y_test, y_pred_svc_sc))
```

```
Confusion matrix of SVC model: 
 [[406   5]
 [ 12 330]] 

Accuracy of SVC model =  0.9774236387782205
```
