## Supplementary material for "Comparative Evaluation Of Machine Learning Classifiers For Brain Tumor Detection": contains: supplemental file, brain_tumor_detection_model.ipynb, brain_tumor_detection_model.html, brain_tumor_dataset.csv: brain_tumor_detection_model.ipynb

```
# %% [code]
import numpy as np  # for numeric calculation
import pandas as pd  # for data analysis and manupulation
import matplotlib.pyplot as plt  # for data visualization
import seaborn as sns  # for data visualization

# %% [code]
# import dataframe
url = 'https://raw.githubusercontent.com/1umairali/models/main/brain_tumor_detection/brain_dataset.csv'
brain_dataframe = pd.read_csv(url)
brain_dataframe

# %% [code]
# Head (first six rows) of brain dataframe
brain_dataframe.head(6)

# %% [code]
# Tail (last six rows) of brain dataframe
brain_dataframe.tail(6)

# %% [code]
# Information of brain dataframe
brain_dataframe.info()

# %% [code]
# show image columns
print(brain_dataframe['Image'])

# %% [markdown]
image column is object dtype. contains only images name. if we drop that column it will not impact dataframe or result

# %% [code]
# drop Image column.
brain_df2 = brain_dataframe.drop(['Image'], axis = 1)
brain_df2

# %% [code]
# Numerical distribution of data
brain_df2.describe()

# %% [code]
# check sum of null values in each columns
brain_df2.isnull().sum()

# %% [markdown]
## Data Visualization

# %% [code]
# Pairplot of brain dataframe
sns.pairplot(brain_df2, hue = 'Class')

# %% [code]
# Count the class columns
# 0 = Non tumor / no cancer
# 1 = Tumor / has cancer
sns.countplot(x=brain_df2["Class"])

# %% [markdown]
## Heatmap

# %% [code]
# heatmap of DataFrame
plt.figure(figsize=(16,9))
sns.heatmap(brain_df2)

# %% [markdown]
## Heatmap of a correlation Matrix

# %% [code]
# correlation matrix
brain_df2.corr()

# %% [markdown]
we checked in information 'Coarseness' column is float datatype. contains 0 values which is not empty.......
but in corelation matrix it contain NaN values......
we have to remove that column. it will not impact the result......
lets confirm in Heatmap and Barplot of Correlation Matrix

# %% [code]
# Heatmap of Correlation matrix of Brain DataFrame
plt.figure(figsize=(10,8))
sns.heatmap(brain_df2.corr(), annot = True, cmap ='coolwarm', linewidths=2)

# %% [code]
# drop coarseness column from brain_df2
brain_df3 = brain_df2.drop(['Coarseness'], axis=1)

# %% [markdown]
## Split Dataframe in Train and Test

# %% [code]
# drop dependent (Class) column, it will assign to y
X = brain_df3.drop(['Class'], axis = 1)
X.head(6)

# %% [code]
# output variable
# assign Class column to y
y = brain_df3['Class']
y.head(6)

# %% [code]
# split dataset into train and test
from sklearn.model_selection import train_test_split
X_train, X_test, y_train, y_test = train_test_split(X, y, test_size = 0.2, random_state= 5)

# %% [code]
X_train

# %% [code]
X_test

# %% [code]
y_train

# %% [code]
y_test

# %% [markdown]
## Feature Scaling

# %% [code]
from sklearn.preprocessing import StandardScaler
sc = StandardScaler()
X_train_sc = sc.fit_transform(X_train)
X_test_sc = sc.transform(X_test)

# %% [markdown]
## ML Model Building

# %% [code]
from sklearn.metrics import confusion_matrix, classification_report, accuracy_score

# %% [markdown]
## Support Vector Classifier

# %% [markdown]
##### Train with original data 

# %% [code]
from sklearn.svm import SVC
svc_classifier = SVC()
svc_classifier.fit(X_train, y_train)
y_pred_svc = svc_classifier.predict(X_test)
accuracy_score(y_test, y_pred_svc)

# %% [markdown]
##### Train with scaled data

# %% [code]
svc_classifier2 = SVC()
svc_classifier2.fit(X_train_sc, y_train)
y_pred_svc_sc = svc_classifier2.predict(X_test_sc)
accuracy_score(y_test, y_pred_svc_sc)

# %% [markdown]
## Logistic Regression

# %% [markdown]
##### Train with original data 

# %% [code]
from sklearn.linear_model import LogisticRegression
lr_classifier = LogisticRegression(random_state=51, penalty = 'l2')
lr_classifier.fit(X_train, y_train)
y_pred_lr = lr_classifier.predict(X_test)
accuracy_score(y_test, y_pred_lr)

# %% [markdown]
##### Train with scaled data 

# %% [code]
lr_classifier2 = LogisticRegression(random_state=51, penalty = 'l2')
lr_classifier2.fit(X_train_sc, y_train)
y_pred_lr_sc = lr_classifier2.predict(X_test_sc)
accuracy_score(y_test, y_pred_lr_sc)

# %% [markdown]
## KNN - K-Nearesr Neighbor Classifier

# %% [markdown]
##### Train with original data

# %% [code]
from sklearn.neighbors import KNeighborsClassifier
knn_classifier = KNeighborsClassifier(n_neighbors = 5, metric = 'minkowski', p = 2)
knn_classifier.fit(X_train, y_train)
y_pred_knn = knn_classifier.predict(X_test)
accuracy_score(y_test, y_pred_knn)

# %% [markdown]
##### Train with scaled data

# %% [code]
knn_classifier2 = KNeighborsClassifier(n_neighbors = 5, metric = 'minkowski', p = 2)
knn_classifier2.fit(X_train_sc, y_train)
y_pred_knn_sc = knn_classifier2.predict(X_test_sc)
accuracy_score(y_test, y_pred_knn_sc)

# %% [markdown]
## Naive Bayes Classifier

# %% [markdown]
##### Train with original data

# %% [code]
from sklearn.naive_bayes import GaussianNB
nb_classifier = GaussianNB()
nb_classifier.fit(X_train, y_train)
y_pred_nb = nb_classifier.predict(X_test)
accuracy_score(y_test, y_pred_nb)

# %% [markdown]
##### Train with scaled data

# %% [code]
nb_classifier2 = GaussianNB()
nb_classifier2.fit(X_train_sc, y_train)
y_pred_nb_sc = nb_classifier2.predict(X_test_sc)
accuracy_score(y_test, y_pred_nb_sc)

# %% [markdown]
## Decision Tree Classifier

# %% [markdown]
##### Train with original data

# %% [code]
from sklearn.tree import DecisionTreeClassifier
dt_classifier = DecisionTreeClassifier(criterion = 'entropy', random_state = 51)
dt_classifier.fit(X_train, y_train)
y_pred_dt = dt_classifier.predict(X_test)
accuracy_score(y_test, y_pred_dt)

# %% [markdown]
##### Train with scaled data

# %% [code]
dt_classifier2 = DecisionTreeClassifier(criterion = 'entropy', random_state = 51)
dt_classifier2.fit(X_train_sc, y_train)
y_pred_dt_sc = dt_classifier2.predict(X_test_sc)
accuracy_score(y_test, y_pred_dt_sc)

# %% [markdown]
## Random Forest Classifier

# %% [markdown]
##### Train with original data

# %% [code]
from sklearn.ensemble import RandomForestClassifier
rf_classifier = RandomForestClassifier(n_estimators = 20, criterion = 'entropy', random_state = 51)
rf_classifier.fit(X_train, y_train)
y_pred_rf = rf_classifier.predict(X_test)
accuracy_score(y_test, y_pred_rf)

# %% [markdown]
##### Train with scaled data

# %% [code]
rf_classifier2 = RandomForestClassifier(n_estimators = 20, criterion = 'entropy', random_state = 51)
rf_classifier2.fit(X_train_sc, y_train)
y_pred_rf_sc = rf_classifier2.predict(X_test_sc)
accuracy_score(y_test, y_pred_rf_sc)

# %% [markdown]
## Confusion Matrix

# %% [code]
cm = confusion_matrix(y_test, y_pred_rf_sc)
plt.title('Heatmap of Confusion Matrix', fontsize = 15)
sns.heatmap(cm, annot = True)
plt.show()

# %% [markdown]
## Classification Report Of model

# %% [code]
print(classification_report(y_test, y_pred_rf_sc))

# %% [markdown]
## Cross-validation of the ML model

# %% [code]
# Cross validation
from sklearn.model_selection import cross_val_score
cross_validation = cross_val_score(estimator = rf_classifier2, X = X_train_sc,y = y_train, cv = 10)
print("Cross validation accuracy of SVC model = ", cross_validation)
print("\nCross validation mean accuracy of SVC model = ", cross_validation.mean())

# %% [markdown]
## Test Model

# %% [code]
# we have one patient data
patient1 = [17.99,10.38,122.8,1001.0,0.1184,0.2776,0.3001,
            0.1471,0.2419,0.07871,1.095,0.9053]

# %% [code]
# convert list into array and scale
patient1_sc = sc.transform(np.array([patient1]))
patient1_sc

# %% [code]
# predict patient1_sc scale data
# zero mean malignant patient has cancer
predict = rf_classifier2.predict(patient1_sc)
predict

# %% [code]
# write if else statement to print result in clear format
if predict[0] == 0:
    print ('Patient has *** NO *** Tumor / Cancer')
else:
    print ('Patient *** HAS *** Tumor / Cancer')

# %% [code]
# confusion matrix
print('Confusion matrix of Random Forest model: \n',confusion_matrix(y_test, y_pred_rf_sc),'\n')

# show the accuracy
print('Accuracy of Random Forest model = ',accuracy_score(y_test, y_pred_rf_sc))
```
