## Supplementary material for "Comparative Evaluation Of Machine Learning Classifiers For Brain Tumor Detection": contains: supplemental file, brain_tumor_detection_model.ipynb, brain_tumor_detection_model.html, brain_tumor_dataset.csv: readme.txt

Supplemental file 1.pdf				is complete detail of manuscript.

brain_tumor_detection_model.ipynb		is Model of detect brain tumor. this file opens in Jupyter Notebook.

brain_tumor_detection_model.html		is converted version of brain_tumor_detection_model.ipynb file. just for viewing and understanding.

brain_tumor_dataset.csv				contains samples data of patients (who has brain tumor, and who hasn't) downloaded from https://www.kaggle.com/datasets/jakeshbohaju/brain-tumor?resource=download
