## Supplementary figures and images for "Comparative Evaluation Of Machine Learning Classifiers For Brain Tumor Detection"

### Fig. 1. MRI images of patients without brain tumor.jpg

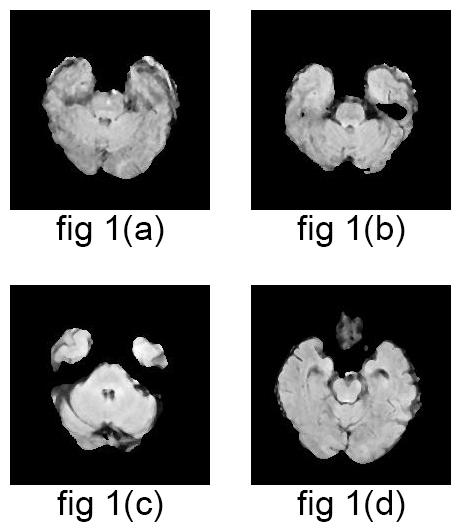

### Fig. 2. MRI images of patients with brain tumor.jpg

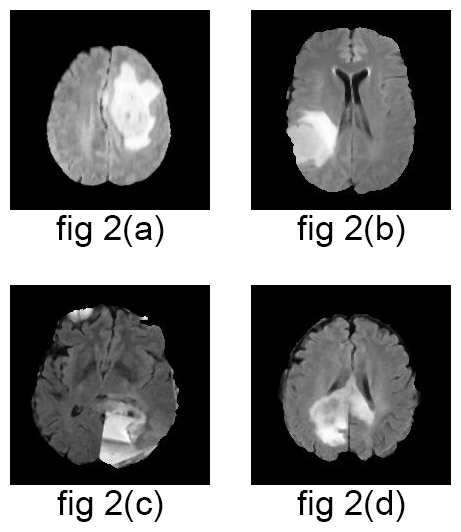
